# Radiotherapy continuity for cancer treatment: lessons learned from natural disasters

**DOI:** 10.1101/2024.07.18.24310636

**Authors:** Ralf Müller-Polyzou, Melanie Reuter-Oppermann

**Author notes:** These authors contributed equally to this work.

## Abstract

**Background:** The contemporary world is challenged by natural disasters accelerated by climate change, affecting a growing world population. Simultaneously, cancer remains a persistent threat as a leading cause of death, killing 10 million people annually. The efficacy of radiotherapy, a cornerstone in cancer treatment worldwide, depends on an uninterrupted course of therapy. However, natural disasters cause significant disruptions to the continuity of radiotherapy services, posing a critical challenge to cancer treatment. This paper explores how natural disasters impact radiotherapy practice, compares them to man-made disasters, and outlines strategies to mitigate adverse effects of natural disasters. Through this analysis, the study seeks to contribute to developing resilient healthcare frameworks capable of sustaining essential cancer treatment amidst the challenges posed by natural disasters.

**Method:** We conducted a Structured Literature Review to investigate this matter comprehensively, gathering and evaluating relevant academic publications. We explored how natural disasters affected radiotherapy practice and examined the experience of radiotherapy centres worldwide in resuming operations after such events. Subsequently, we validated and extended our research findings through a global online survey involving radiotherapy professionals.

**Results:** The Structured Literature Review identified twelve academic publications describing hurricanes, floods, and earthquakes as the primary disruptors of radiotherapy practice. The analysis confirms and complements risk mitigation themes identified in our previous research, which focused on the continuity of radiotherapy practice during the COVID-19 pandemic. Our work describes nine overarching themes, forming the basis for a taxonomy of 36 distinct groups. The subsequent confirmative online survey supported and solidified our findings and served as a basis for developing a conceptual framework for natural disaster-resilient radiotherapy.

**Discussion:** The growing threat posed by natural disasters underscores the need to develop business continuity programs and define risk mitigation measures to ensure the uninterrupted provision of radiotherapy services. By drawing lessons from past disasters, we can better prepare for future hazards, supporting disaster management and planning efforts, particularly enhancing the resilience of radiotherapy practice. Additionally, our study can serve as a resource for shaping policy initiatives aimed at mitigating the impact of natural hazards.

## Introduction

Global climate change and its effects are ubiquitous. Greenhouse gas emissions remain at unprecedented levels, and the year 2023 was reported to be one of the warmest in historical records. High water temperatures in the ocean and the loss of glacier mass led to rising sea levels. At the same time, severe weather conditions were reported, such as heatwaves, droughts, tropical cyclones, and floodings [1]. In total, 399 natural hazards and disasters resulted in 86,473 deaths, impacting 93.1 million people and causing economic losses of USD 202.7 billion in 2023 [2]. These figures impressively reflect the importance of preparing for future natural disasters, also considering that two-thirds of all disasters worldwide since 2000 have been related to natural hazards [3]. Despite the efforts made, only a few years remain for The Sendai Framework for Disaster Risk Reduction 2015-2030 to fulfil its promises to prevent and reduce hazard exposure and vulnerability to disaster, increase preparedness for response and recovery, and strengthen resilience [4]. Climate change, population growth, and weather sensitivities demand early warning and forecasting skills that help to avoid the damage caused by extreme weather events. This is especially important for Least Developed Countries (LDC) and Small-Island Developing States (SIDS) that are hit hard by the impact of global climate change [5].

The 2022 report of the Lancet Countdown on health and climate change outlines the effect of natural disasters on global health, stating that: “[…] extreme weather events are increasingly affecting physical and mental health directly and indirectly […]” [6]. Patients with noncommunicable diseases, including cancer, possess unique needs and high vulnerability to treatment disruptions caused by natural disasters. Risk reduction strategies must, therefore, incorporate their specific needs throughout all stages of disaster management [7]. However, most articles on cancer care during and after natural disasters use a general approach [8]. They do not consider the specifics of individual therapy forms, such as radiotherapy (RT), a treatment modality delivered in multiple strictly consecutive sessions, often referred to as treatment fractions, to minimise patient side effects.

As climate change intensifies, natural disasters become more prevalent. Consequently, systems, tools, and policies are required to mitigate the risks to RT effectively. Developing such solutions demands interdisciplinary research and a meaningful consolidation of best practices from past experiences. To support this, we have reviewed academic literature and conducted an online survey on RT practice in the context of natural disasters. The main contributions of this work are (1) a collection of academic publications about RT disaster management, the resilience of RT practice, and rebuilding efforts following natural disasters, (2) a content analysis of the identified literature supporting the mitigating of the impact that future natural disasters can have on RT practice, (3) results of a confirmative online survey on business continuity risks and risk mitigation measures for RT, and (4) a conceptual framework for natural disaster resilient RT. The paper builds on previous work of our research group, expanding the knowledge base of Business Continuity Management (BCM) in RT practice [9].

The paper is structured as follows: First, important foundations are laid. Natural hazards and disasters are introduced, and the link between increasing cancer incidence and climate epidemiology is established. Treatment with RT is described as an important form of cancer therapy, and the effect of natural hazard-induced treatment interruptions is presented. Furthermore, BCM and Risk Management (RM) are outlined, followed by the methodological background of the Systematic Literature Review (SLR) and the structured online survey. We then present the data analysis concept, the results obtained and the conceptual model. The results are discussed afterwards, followed by the conclusions and outline of topics for future research. Supporting information is provided through a literature summary in S1, coding data in S2, survey data in S3 and the structured online questionnaire in S4.

## Foundations

### Natural hazards and disasters

A natural hazard encompasses geophysical processes inherent to the environment, presenting risks to human lives and property. Natural hazards comprise climate- and meteorology-related geophysical phenomena and those originating from geology and geomorphology, which are often combined with each other. Natural hazards can be categorised according to their cause as shown in Table 1. Natural hazards become natural disasters when their consequences result in substantial loss of life and property, surpassing the capacity of local communities to recover independently. In this context, global population growth, urbanisation, and coastalisation increase the risk of the occurrence of natural disasters [10–12]. Numerous academic publications focus on the impact of natural disasters on healthcare, health impact and cancer, particularly [13–15]. The American Cancer Society even published rules for emergencies, advising patients to create a list of medications and treatment schedules and obtain additional medication supplies. The society also recommends keeping a record of important phone numbers, discussing options for rescheduling treatment, and getting vaccinated against potential diseases [16]. However, these rules are not RT-specific and address only actions under patients’ responsibility.

**Table 1.**
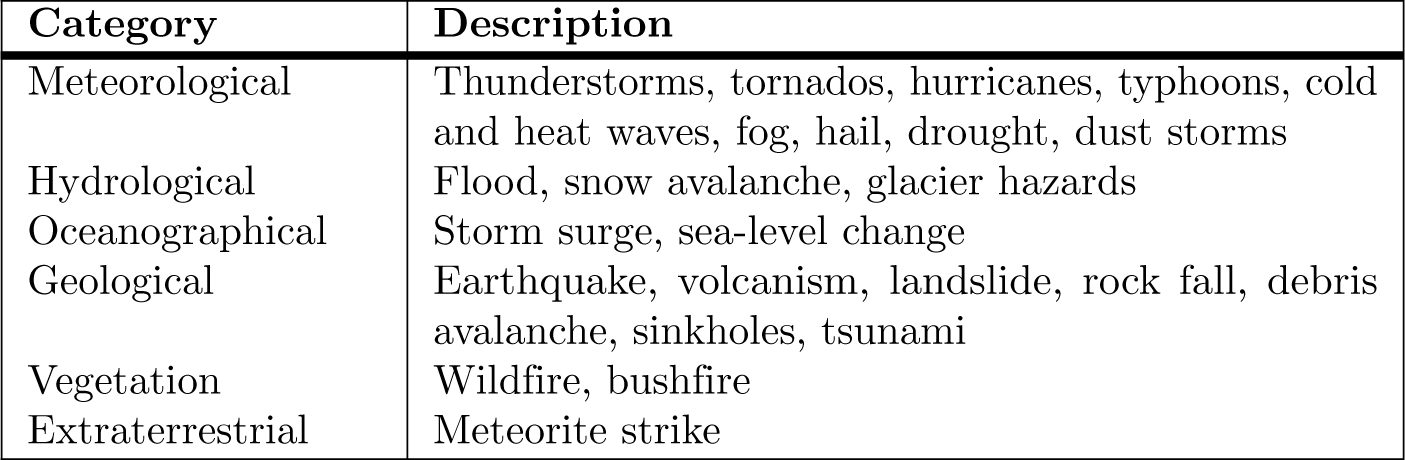
Categorisation of natural hazards.

### Cancer and radiotherapy

Cancer is the second leading cause of global mortality, accounting for approximately 10 million deaths every year [17]. Cancer patients will surge by 75 % in the next two decades due to ageing populations and unhealthy lifestyles [18]. Studies suggest that around 50 % of cancer patients worldwide could benefit from RT [19]. Treatment with RT is a therapy that constitutes a technologically driven domain within healthcare. Diagnostic imaging techniques, including Computed Tomography, Positron Emission Tomography Computed Tomography, and Magnetic Resonance Imaging, play a crucial role in diagnosis and treatment planning, while Linear Accelerators (LINACs) precisely deliver the radiation dose to the tumour. Modern RT relies on integrated information systems operated by specialised staff who interact with patients and colleagues [20]. Advanced planning systems calculate individualised treatment plans, while quality assurance assessments are conducted to verify these plans. Given the interconnectivity of RT systems, extensive data processing occurs, mainly in the form of large image files. As LINACs have emerged, the technology has evolved to encompass complex irradiation techniques.

### Business continuity and risk management

Attempts to ensure the uninterrupted provision of business services are described in BCM. The ISO 22,301 standard delineates the requirements for implementing, maintaining, and enhancing a robust BCM system. Conversely, RM encompasses activities pertaining to identifying, analysing, evaluating, controlling, and monitoring risks. A well-defined BCM plan is valuable in effectively managing risks [21]. While technical redundancy holds importance in many BCM projects, it alone does not suffice to mitigate risks comprehensively, as many have experienced during the COVID-19 pandemic. BCM, consequently, encompasses people and interdependencies with suppliers, co-workers, partners, patients, and even their families. All of these are parameters particularly valid for RT practice. Subsequently, risk strategies face many challenges natural disasters pose, while BCM and support systems are essential for safe and efficient RT services in adverse conditions.

### Systematic Literature Review

An SLR analyses the knowledge base by applying a transparent, reproducible, and unbiased selection and review of the literature. The selected articles are synthesised through content analysis, creating knowledge [22, 23]. We compiled literature using configurative review incorporating inductive interpretation and exploration. Initially, we formulated a search strategy encompassing the breadth, depth, time, resources, and inclusion and exclusion criteria as documented in the search protocol shown in Table 2. The search terms were defined based on the index of a textbook of radiation physics [24]. To mitigate bias, we included synonyms and variations in the spelling of terms. The search process was conducted using multiple academic databases with forward and backward searches. The selection took place based on the abstracts. Articles, review articles, editorials, and book chapters published in English between January 2000 and May 2023 were considered. The literature management software Citavi was used for screening and sorting the literature [25]. An additional search in Scopus for the adjacent BCM and RM areas did not add further literature. A complementary search covering June 2023 to April 2024 identified 86 articles but did not add relevant ones. After identifying relevant studies, duplicates were removed, and a broad screening process was conducted. Selected articles underwent a full-text assessment, and a final set was content analysed using manual coding techniques and categorisation. To ensure transparency, we followed the guidelines outlined in the 2009 edition of the Preferred Reporting Items for Systematic Reviews and Meta-Analysis (PRISMA) [26].

**Table 2.** Search protocol of the SLR.

| Section | Description |
| --- | --- |
| Conceptual frame | Cancer as a global non-communicable disease |
| Context | Radiotherapy treatment following disruptions caused by natural hazards and disasters |
| Period | January 2000 to May 2023, complementary June 2023 to April 2024 |
| Language | English |
| Strategy | Aggregating search |
| Focus | Title, abstract, keywords |
| Literature | Articles, review articles, editorials, book chapters |
| Sources | Scopus, ScienceDirect, Web of Science, Google Scholar |
| Search string Scopus | (radiotherapy OR "radiation therapy" OR teletherapy) AND (hazard OR disaster OR risk OR threat OR danger) AND (nature OR climate) |
| Categories included Scopus | Medicine, physics and astronomy, health professions, nursing, environmental science, engineering, computer science |
| Search string ScienceDirect | (radiotherapy OR "radiation therapy") AND (hazard OR disaster OR risk OR threat OR danger) AND (nature OR climate) |
| Categories included ScienceDirect | All subject areas |
| Search string Web of Science | (radiotherapy OR "radiation therapy" OR teletherapy) AND (hazard OR disaster OR risk OR threat OR danger) AND (nature OR climate) |
| Categories included Web of Science | All subject areas |
| Search string Google Scholar | (radiotherapy OR "radiation therapy") AND (disaster) AND (nature OR climate) |
| Categories included Google Scholar | All subject areas |
| Search Strategy Google Scholar | Full text, all articles sorted by relevance, first 200 screened by title. After the first 50 results, the relevance dropped, but papers were still found to be eligible. In total, 200 papers were screened. |
| Inclusion criteria | External beam radiotherapy; natural hazard; disaster |
| Exclusion criteria | No relation to radiotherapy; clinical trials; clinical studies; human-caused disasters |

### Structured online survey

We implemented a structured online survey using Google Forms to validate and extend our research findings involving RT professionals [27]. The questionnaire was reviewed by two RT experts to increase the probability of acceptance and participation. Participants were introduced to the survey’s scientific background, informed about the time required for completion, and assured of the confidentiality of their responses. Most survey questions were structured as closed-ended or semi-open inquiries to promote objectivity. A 5-point unipolar Likert scale with and without an ”I don’t know” option was thoughtfully employed to gauge participant sentiment regarding specific statements, fostering quantifiable analysis. The results were analysed using Top-2-Box (T2B) agreement and Buttom-2-Box (B2B) disagreement values. The interpretation of Likert scores depends on the study’s context and the research’s specific objectives. We consider approval ratings of 80 % and above and T2B to B2B ratios of ten or more unambiguous [28]. Before finalising their responses, participants were encouraged to share additional comments and suggestions. We integrated the structured online survey with the SLR to establish a robust foundation for our research.

## Data analysis

The SLR, content analysis, and survey results are depicted in the following subsections, supported by literature summaries, coding and survey data, and the questionnaire documented in the supporting information S1 to S4.

### Results of the SLR

A total of 1691 articles published from January 2000 to April 2024 have been identified, following the removal of duplicate articles using the literature management tool. Afterwards, 1,674 records were excluded based on exclusion parameters. Subsequently, 17 articles remained, which underwent further analysis. Five more articles were excluded during the full-text assessment. The structure of the SLR is shown in Fig 1, presenting the PRISMA chart, illustrating the stages of the identification and screening process, along with the number of articles handled at each step. The included literature is presented in Table 3 structured by author, year of publication, title, and the kind of natural hazard reflected in the article.

**Fig 1.**
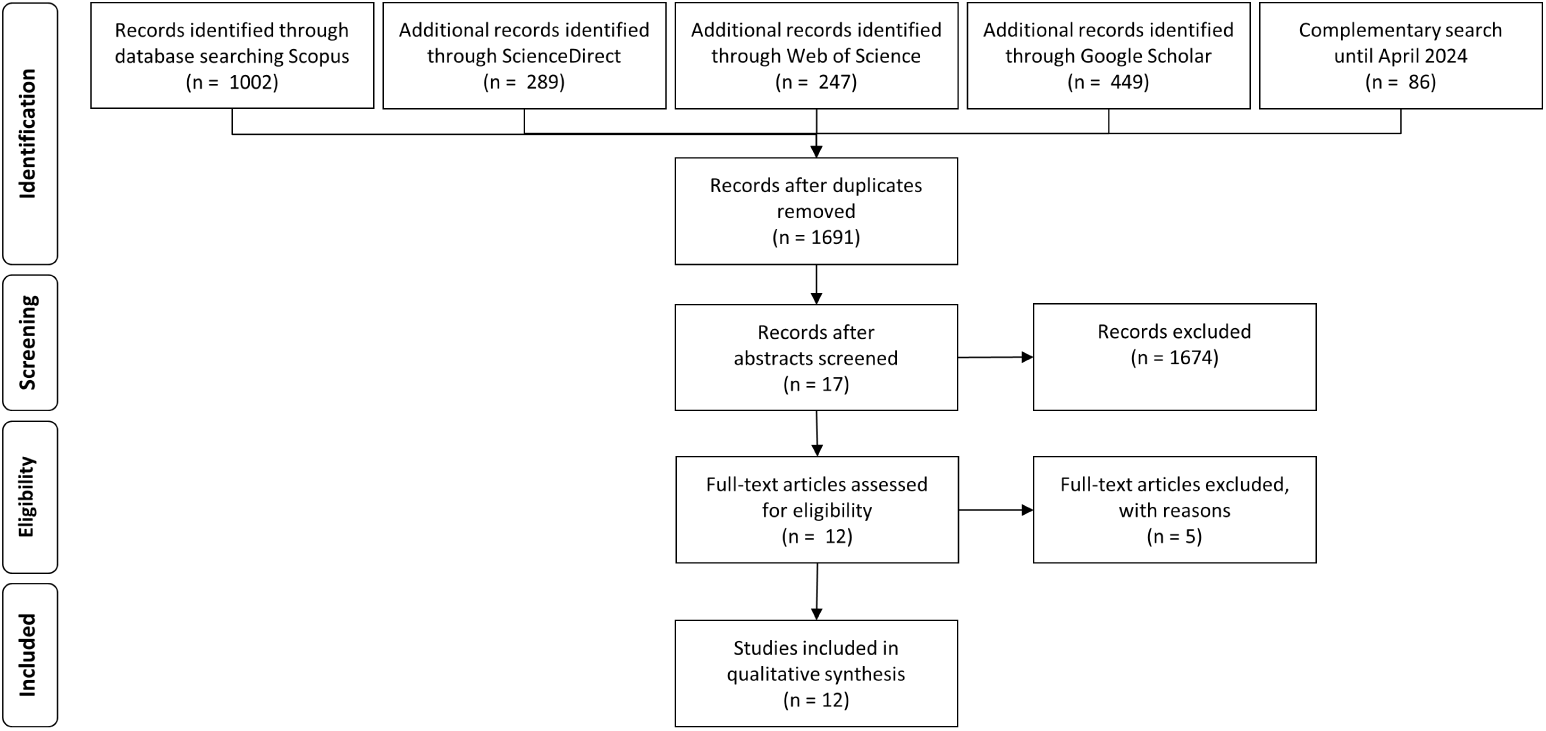
PRISMA chart of the SLR.

**Table 3.** Literature included in the qualitative synthesis.

| Author | Year | Title | Hazard |
| --- | --- | --- | --- |
| Anacak et al. [29] | 2023 | Radiotherapy facilities after the Türkiye–Syria earthquakes: lessons from the tragedy | Earthquake |
| Espinel et al. [30] | 2022 | Climate-driven Atlantic hurricanes create complex challenges for cancer care. | Hurricane |
| Gay et al. [31] | 2019 | Lessons Learned From Hurricane Maria in Puerto Rico: Practical Measures to Mitigate the Impact of a Catastrophic Natural Disaster on Radiation Oncology Patients | Hurricane |
| Grew et al. [32] | 2013 | The Impact of Superstorm Sandy on the Care of Radiation Oncology Patients | Storm |
| Joob and Wiwanitkit [33] | 2012 | Lesson for management of cancerous patient in the big flooding | Flood |
| Lopez-Araujo et al. [34] | 2017 | Letter from Puerto Rico: The State of Radiation Oncology After Maria’s Landfall | Hurricane |
| Man et al. [35] | 2018 | The effect of natural disasters on cancer care: a systematic review. | General |
| Mireles et al. [36] | 2018 | Radiation Oncology in the Face of Natural Disaster: The Experience of Houston Methodist Hospital | Tropical storms, hurricanes |
| Ozaki and Tsubokura [37] | 2018 | Radiation Oncology and Related Oncology Fields in the Face of the 2011 “Triple Disaster” in Fukushima, Japan | Earthquake, tsunami, nuclear meltdown |
| Pérez-Andújar [38] | 2018 | Puerto Rico: After María. | Hurricane |
| Roach et al. [39] | 2018 | Natural Disasters and the Importance of Minimizing Subsequent Radiation Therapy Interruptions for Locally Advanced Lung Cancer. | General |
| Royce et al. [40] | 2019 | Carolina Hurricanes. | Hurricane |

Six articles identified in the SLR describe the impact of hurricanes, which include a combination of meteorological, hydrological, and oceanographical hazards (see Table 1) and can thus quickly develop into comprehensive natural disasters with a high impact on RT practice. Furthermore, earthquakes, which fall into the category of geological hazards, are described. In contrast to hurricanes, there is usually no warning time for earthquakes. Hurricanes, floods, and earthquakes are the natural disasters that have caused the greatest economic damage as a share of Gross Domestic Product on a worldwide decade average since 1960 [41]. Most publications describe disasters in the USA, which are regularly hit by hurricanes but have also established a strong BCM and safety culture.

### Content analysis

The selected articles were manually coded using a mixed coding approach. Codes were assigned to risk mitigation measures described in the identified literature. The codes were first clustered into eight themes: *organisation*, *collaboration*, *communication*, *access*, *protection*, *therapy*, *facility*, and *data*. Afterwards, codes were categorised into groups. Themes developed in previous research, as shown in Fig 2, were deductively used as a basis for code assignment and connect our work with previous research focusing on RT during the COVID-19 pandemic [9]. Two new themes, *facility* and *data*, were developed inductively from the identified literature. New code groups enhanced the existing themes, while the *hygiene* theme from our pandemic research was not used. The new code groups are highlighted in grey in Fig 2. The reliability of the content analysis was increased by a review of the coding process and results.

**Fig 2.**
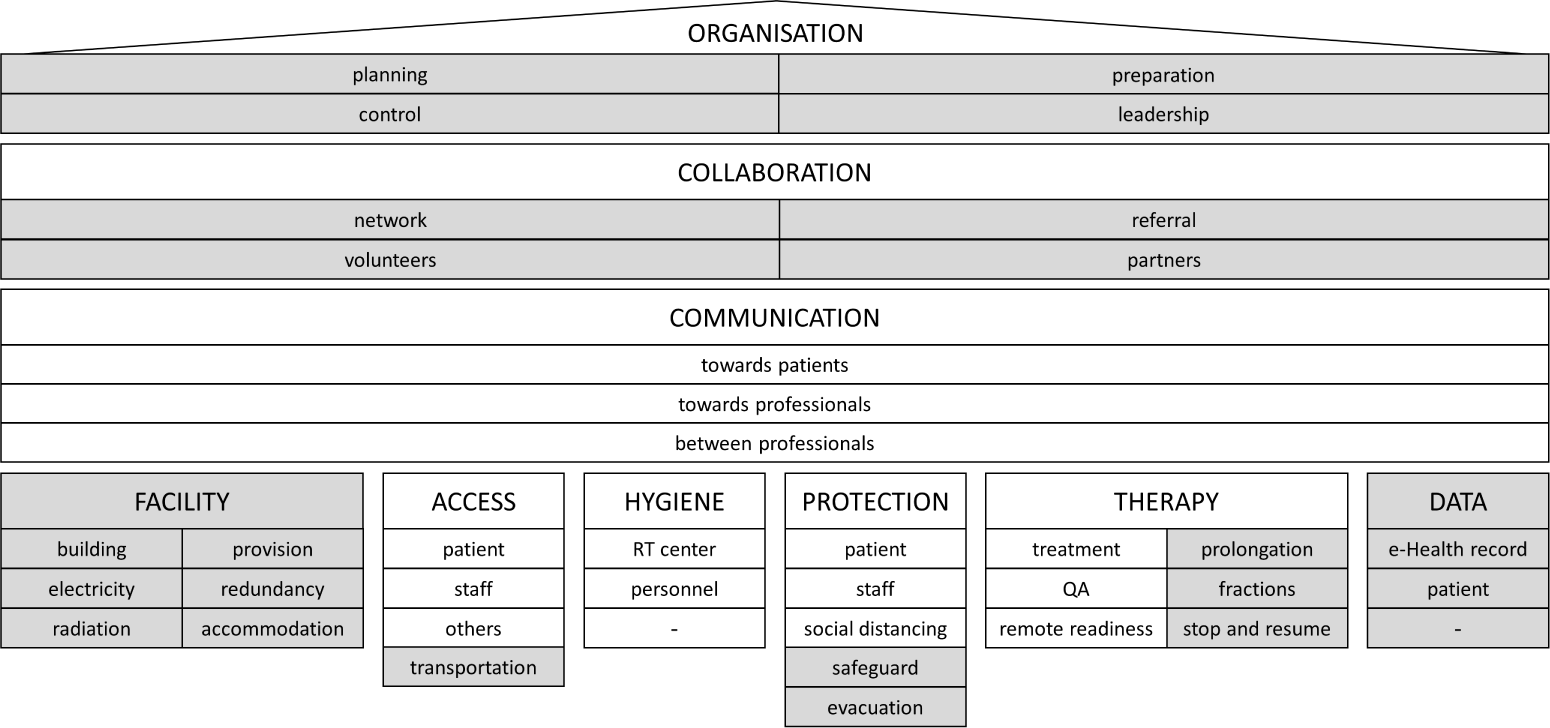
Taxonomy of risk mitigation measures.

The supporting information S2 contains the themes, code groups, description, and code frequency. Eight themes and 25 code groups were used based on 79 codes assigned during the content analysis. The number of code groups, codes and the code frequency per code group of the identified themes are shown in Table 4. The distribution shows that the first three themes dominate the analysis with cumulative 62 % of all codes and *collaboration* being the theme with the highest code frequency. The analysis of code co-occurrence did not provide additional information. With the additions to the taxonomy presented in [10], the taxonomy now consists of nine themes and 36 groups, as shown in Fig 2.

**Table 4.** Quantitative presentation of codes and code groups of themes.

| Themes | Code Groups | Codes | Frequency | Frequency cum. |
| --- | --- | --- | --- | --- |
| Collaboration | 4 | 22 | 27.8 | 27.8 |
| Organisation | 4 | 15 | 19.0 | 46.8 |
| Therapy | 3 | 12 | 15.2 | 62.0 |
| Communication | 3 | 9 | 11.4 | 73.4 |
| Data | 2 | 9 | 11.4 | 84.8 |
| Facility | 6 | 7 | 8.9 | 93.7 |
| Protection | 2 | 3 | 3.8 | 97.5 |
| Access | 1 | 2 | 2.5 | 100.0 |
| **Total** | **25** | **79** | **100.0** | **100.0** |

Summarising, the theme *organisation* includes governance and staff management activities that are critically important in crisis situations. The underlying *collaboration* theme reflects dependencies on internal and external suppliers and cooperation with co-workers, partners and volunteers. Internal and external *communication* is fundamental for risk mitigation. The theme includes *communication* between patients and professionals as well as among professionals of RT centres. The foundations are the themes *facility*, *access*, *hygiene*, *protection*, *therapy* and *data*. The themes *access*, *hygiene*, *protection*, and *therapy* include measures for the management of infection-related crises. *Access* measures are applied to patients, staff, and others who want to enter RT centres and now also include transportation aspects. *Hygiene* measures are important to prevent infections and are applied during pandemics. *Protection* for patients and staff with Personal Protection Equipment is known in healthcare environments. Social distancing adds an additional level of protection. Safeguarding and evacuation were added, reflecting natural disaster needs. *Therapy* is the core of RT cancer treatment and was extended with additional measures increasing treatment flexibility. The themes *facility* and *data* add dimensions for natural disaster-specific mitigation measures.

### Results of the survey

#### Sample group and implementation

The structured online survey addressed the professional groups working in RT centres to consider multiple perspectives. The survey in English was accessible from November 13, 2023, to January 31, 2024, and was promoted via email, the LinkedIn professional network, and Facebook social media. During this time, industry leaders encouraged survey participation by commenting on and reposting the call for participation. In total, 111 participants fully completed the survey and were thereby eligible for analysis. The high response rate can be attributed to a motivating personal address, our personal donation for cancer aid, and concise questions with varying answer formats.

The participants came from the following countries: Germany (56), Ukraine (20), Greece (5), United Kingdom (5), United States (4), Austria (3), Netherlands (3), and one each from Bangladesh, Egypt, Grenada, Hungary, Indonesia, Iran, Republic of Ireland, New Zealand, Oman, Serbia, Singapore, South Africa, St Kitts & Nevis, Switzerland and Turkey. The sample group consists of participants from private and public hospitals and RT centres, as shown in Table 5. Mainly, medical physicists and dosimetrists took part in the survey (82), but also radiation oncologists (15), radiation therapists (12), administrators (1), and others (1). Most participants from Germany were medical physicists (n=54), while the participants from Ukraine included 50 % radiation oncologists (10).

**Table 5.**
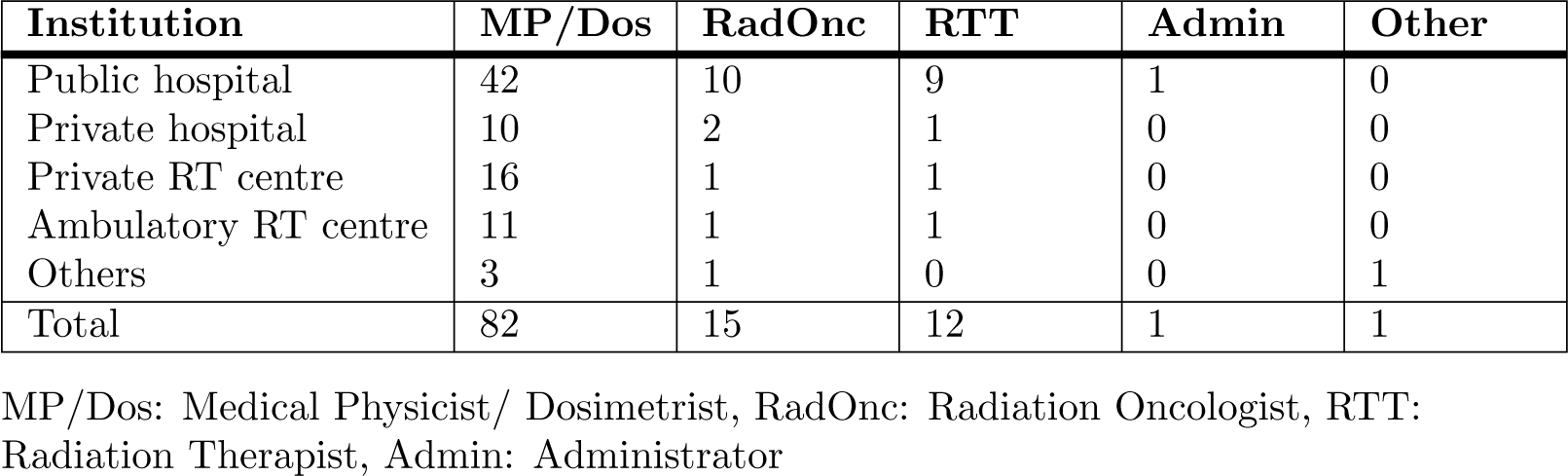
Structure of the sample group (n=111).

Overall, we can recognise a predominance of German and Ukrainian participants, likely since the questionnaire was additionally promoted via email in those countries. Also, the medical physicist professional group predominates, probably because of the existing social media connections. The distribution between the different types of institutions is balanced, with almost 70 % of institutions located in multi-storey buildings, as shown in Table 6. Five participants claim they do not work in RT centres. Considering the special context of this survey, these participants could come from governmental institutions, associations or private companies. Overall, the participants are a solid, but not representative, sample group for the empirical analysis.

**Table 6.** Building type (n=111, single selection).

| Institution | MF | MF-P | SF | SF-P | Other |
| --- | --- | --- | --- | --- | --- |
| Public hospital | 44 | 39.6 | 17 | 15.3 | 1 |
| Private hospital | 12 | 10.8 | 1 | 0.9 | 0 |
| Private RT centre | 12 | 10.8 | 5 | 4.5 | 1 |
| Ambulatory RT centre | 9 | 8.1 | 4 | 3.6 | 0 |
| Other | 0 | 0 | 4 | 3.6 | 1 |
| Total | 77 | 69.4 | 31 | 27.9 | 3 |

MF: Multi-Floor, MF-P: Multi-Floor per cent, SF: Single-Floor, SF-P: Single-Floor per cent

#### Empiric data

First, to understand the assessment of risks to business activities in general, the participants were asked which natural hazards pose the biggest threat in their region considering a two-year and ten-year timeframe. Tables 7 and 8 show the number of responses, frequency and cumulative frequency of responses. Overall, the participants see an increase in risks of natural hazards. They rate all threats by natural hazards higher in the ten-year period than in the two-year period. The three categories *Cold and heat waves, fog, hail, drought, dust storm*, *Flood* and *Thunderstorm, tornado, hurricane, typhoon* are the highest rated in the two-year and ten-year time frame, considering the 80 % threshold of the cumulative frequency of responses. However, it is remarkable that a third of the participants (n=37) see no direct risk in the 2-year period, but this number decreases for the 10-year period (n=27). It appears that the participants changed their long-term risk perception.

**Table 7.**
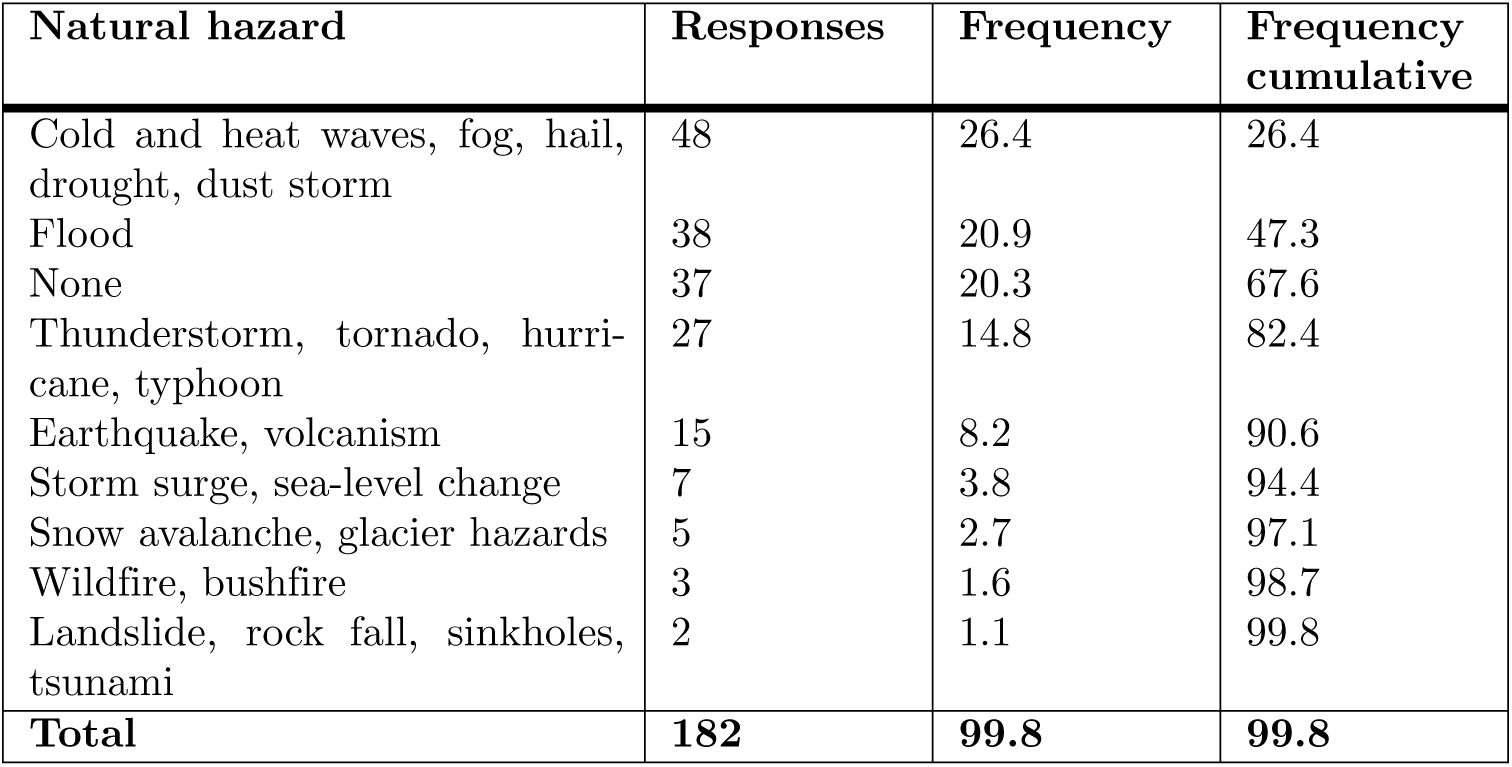
Threats to business activities by natural hazards in a two-year timeframe (n=111, multiple selections possible, except *None*).

| Natural hazard | Responses | Frequency | Frequency cumulative |
| --- | --- | --- | --- |
| Cold and heat waves, fog, hail, drought, dust storm | 48 | 26.4 | 26.4 |
| Flood | 38 | 20.9 | 47.3 |
| None | 37 | 20.3 | 67.6 |
| Thunderstorm, tornado, hurricane, typhoon | 27 | 14.8 | 82.4 |
| Earthquake, volcanism | 15 | 8.2 | 90.6 |
| Storm surge, sea-level change | 7 | 3.8 | 94.4 |
| Snow avalanche, glacier hazards | 5 | 2.7 | 97.1 |
| Wildfire, bushfire | 3 | 1.6 | 98.7 |
| Landslide, rock fall, sinkholes, tsunami | 2 | 1.1 | 99.8 |
| **Total** | **182** | **99.8** | **99.8** |

**Table 8.** Threats to business activities by natural hazards in a ten-year timeframe (n=111, multiple selection possible (except *None*)).

| Natural hazard | Responses | Frequency | Frequency cumulative |
| --- | --- | --- | --- |
| Cold and heat waves, fog, hail, drought, dust storm | 57 | 28.5 | 28.5 |
| Flood | 45 | 22.5 | 51.0 |
| Thunderstorm, tornado, hurricane, typhoon | 31 | 15.5 | 66.5 |
| None | 27 | 13.5 | 80.0 |
| Earthquake, volcanism | 17 | 8.5 | 88.5 |
| Storm surge, sea-level change | 10 | 5.0 | 93.5 |
| Snow avalanche, glacier hazards | 6 | 3.0 | 96.5 |
| Wildfire, bushfire | 4 | 2.0 | 98.5 |
| Landslide, rock fall, sinkholes, tsunami | 3 | 1.5 | 100.0 |
| **Total** | **200** | **100.0** | **100.0** |

We then wanted to understand the risk assessment of natural hazards compared to man-made risks. The participants were asked which risks they perceived as greater in their country than those from natural hazards. The results are shown in Fig 3. In this context, 84.7 % (T2B) of participants rate *cyberattack* as a greater risk than natural hazards. Many of these participants (53.2 %) even strongly agreed with this statement. The risks from *war or terrorism* and *political instability* are still categorised higher by 38.7 % (T2B) and 36.9 % (T2B) of participants, respectively. Considering the continuing war, it is not surprising that the Ukrainian participants see *war and terrorism* (100 %) and *political instability* (95 %) as much higher risks than natural disasters.

**Fig 3.**
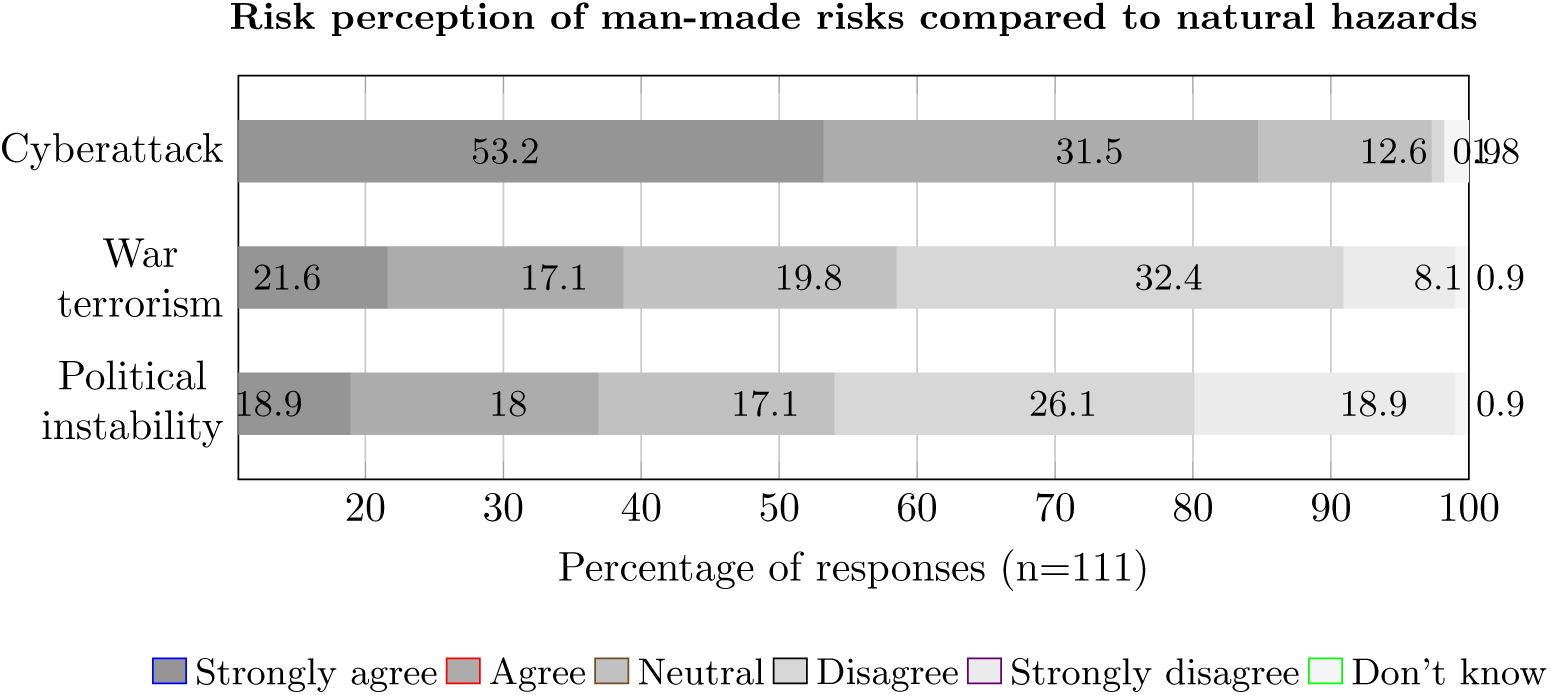
Perceived risks in comparison.

Subsequently, we aimed to understand how well the RT centres are prepared for the risks of natural and man-made hazards. The focus was therefore changed from the general risk situation in the country to the specific RT centre. To do this, we first asked the survey participants whether, in their perception, they knew the risks of the different natural hazards for their RT centre. The results are shown in Fig 4 in descending order of T2B agreement values. The results are indifferent, with relatively few extreme positive or negative ratings. The highest T2B values were noted for *Flood*, *Cold and heat waves, fog, hail, drought, dust storm* and *Thunderstorm, tornado, hurricane, typhoon*, which were also categorised as the biggest hazards in the 2 and 10-year periods. At the same time, 41.4 % (T2B) of participants believe they know the risk of floods to their RT centre.

**Fig 4.**
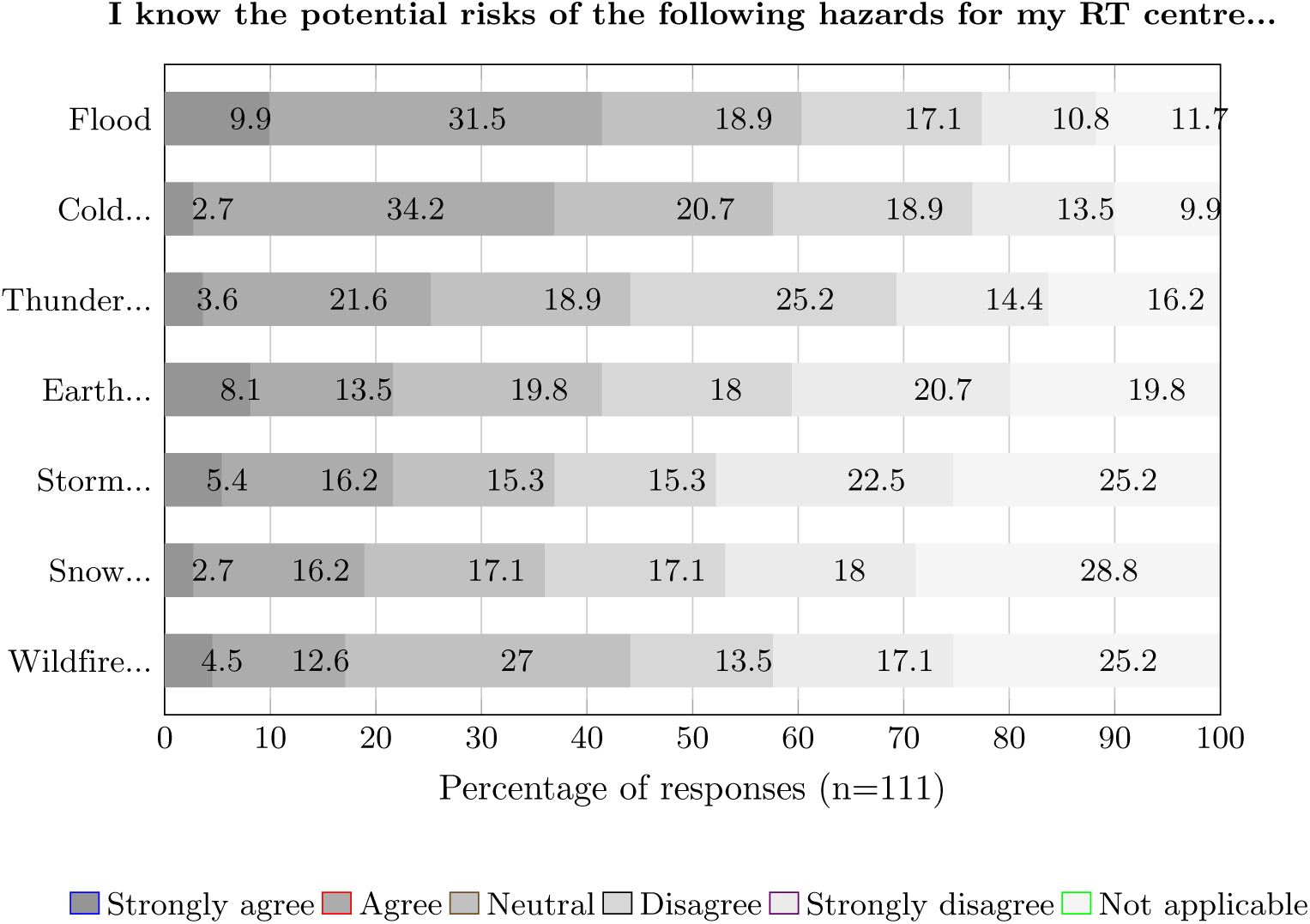
Perception claiming to know the risks of hazards.

We then proceeded to understand how the participants assessed the preparedness of the RT centre, where they work against natural and specific man-made disasters. To do this, we asked whether the participants believed their RT centre was adequately prepared to manage individual risks. The results are shown in Fig 5. The comparison shows that participants consider themselves better prepared against *cyberattacks* (T2B 38.7 %) and *natural hazards* (T2B 37.8 %) than against *political instability* (T2B 19.8 %) or even *war and terrorism*. The participants do not see themselves being prepared against *war and terrorism* in particular (B2B 47.7 %). Overall, the number of neutral responses is high in all four categories. It is particularly striking that in the category of *cyberattacks*, which is considered by far the greatest threat, many participants responded neutrally (30.6 %) or even negatively (B2B 28.8 %). Thus, there is a large discrepancy in preparedness perception. The high number of neutral responses could indicate a lack of BCM strategy with accompanying training and communication in general.

**Fig 5.**
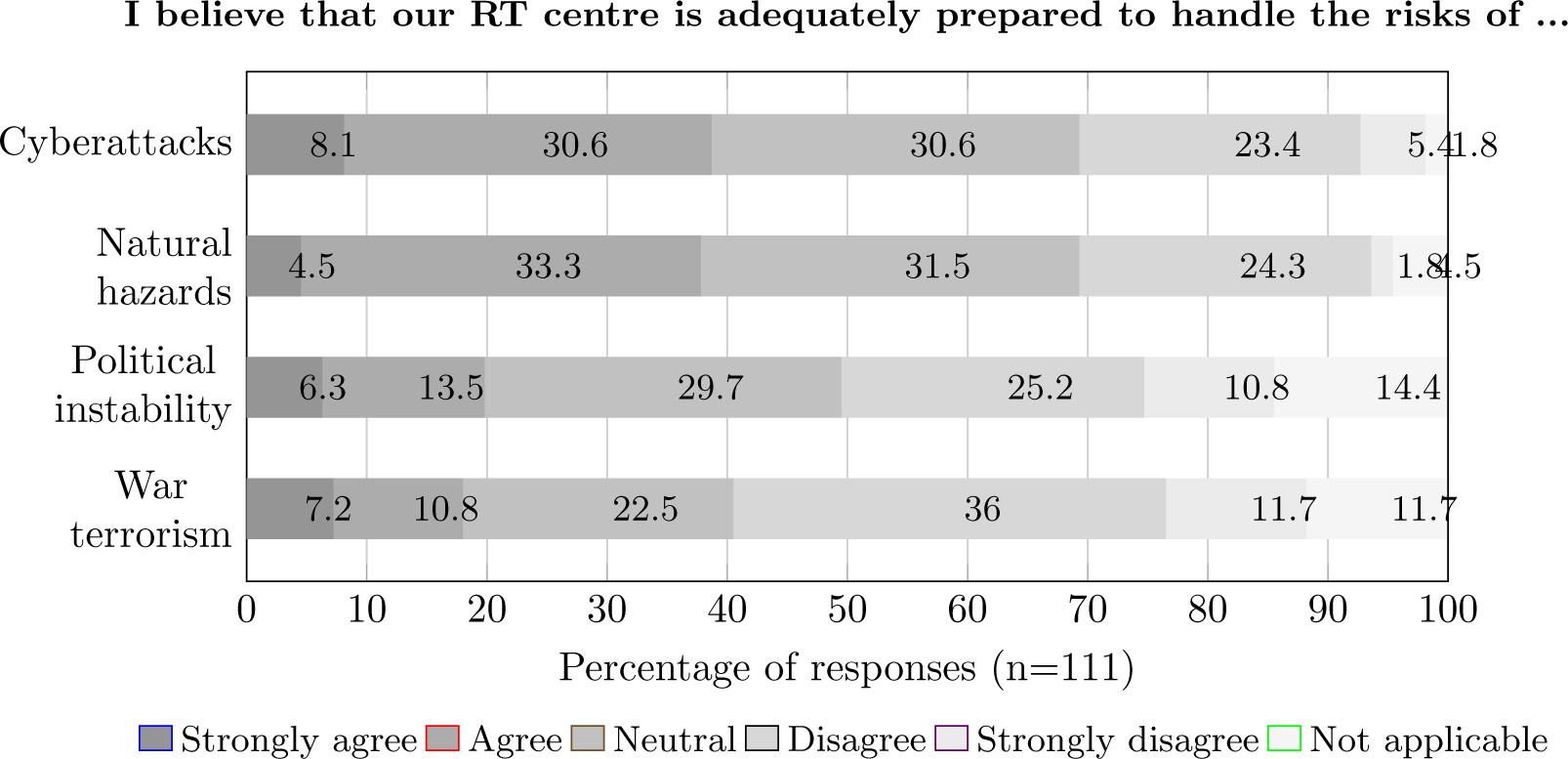
Perception of being able to handle risks.

Following the analysis of the risk awareness and preparedness perception of the sample group, the risk mitigation measures derived from the SLR were rated according to their importance in the categories *organisational*, *communication*, *access, protection, therapy*, *facility* and *data* measures. The supporting information S3 shows the results in descending order of T2B agreement values.

The analysis of the agreement values confirms the identified risk-mitigation measures, often with a high level of agreement, as shown in Table 9 in an aggregated form with the T2B to B2B values ratio, and ten of them being above the defined ratio threshold of ten.

**Table 9.** Aggregated comparison of risk mitigation measures.

| Risk mitigation measures | Ratio |
| --- | --- |
| Transferring patients to cooperating radiotherapy centres to continue treatment based on shared health records and considering offering housing and interpretation services is ... | **19.2** |
| Creating a collaborative network of radiotherapy centres with regional task forces and healthcare coalitions is ... | 9.6 |
| Engaging with vendors, partners, and insurance companies to support emergency offers is ... | 8.1 |
| Organise with the help of radiotherapy associations volunteer staff and aid workers to support or replace own staff and consider special care teams to support patients at home is ... | 3.4 |
| Securing backup communication for leadership and emergency plan execution and preparing for psychological support for caregivers while maintaining an overall positive attitude is ... | **15.8** |
| Securing backup communication methods for communication between caregivers is ... | **18.6** |
| Considering alternative communication methods towards patients, such as Social Media and information provision via the Internet or radio, and preparing for psychological support in patient communication is ... | 7.7 |
| Protecting patients and staff by executing prepared evacuation plans is ... | **17.0** |
| Pausing scheduled radiotherapy treatments and conducting quality assurances of medical devices before resuming treatments is ... | **24.8** |
| Assuring patient's safety and acute needs and safeguarding inpatients to ensure uninterrupted access to care is ... | **15.7** |
| Compensating additional treatment fractions or using hypofractionation techniques to reduce the number of treatment fractions is ... | **12.6** |
| Offering free transportation for patients to radiotherapy centres, utilising reserved fuel stocks is ... | 5.2 |
| Extending the centre's operation hours to evenings and weekends is ... | 2.0 |
| Securing and shutting down sensitive equipment and materials in a controlled manner and protecting against harm, particularly by potential flooding, is ... | **25.3** |
| To twin linear accelerators to minimise the need to recalculate treatment plans in case of device loss is ... | **24.5** |
| Securing electricity supply with an emergency generator, not shared with others, with sufficient fuel storage, and protected against direct and indirect disaster impact is ... | 8.0 |
| Using Electronic Health Records (EHR) with tested recoverable backups in an online repository and data links to cooperating radiotherapy centres is ... | **11.4** |
| Offering accommodation for those unable to commute or being evacuated is ... | 7.9 |
| Providing the patient with updated records during their treatment course is ... | 7.5 |
| Evaluating alternatives for displaced bunker doors securing radiation protection is ... | 4.0 |
| Housing radiotherapy centres in one-floor buildings following strict building codes instead of placing them in multistorey hospital buildings are... | 0.8 |

Ratio: Ratio of T2B to B2B values

Finally, the participants had the opportunity to add comments or suggestions. Overall, it seems that the COVID-19 pandemic raised awareness of potential business disruptions. However, two participants emphasised that the dominant problem remains the lack of qualified staff, a problem that could be addressed using AI technologies and robotic systems. In addition, automatic entrance doors were recognised as a specific solution for access control. Statements such as ”There have never been any natural disasters here, but even small thunderstorms can lead to power outages […]” ”We experience intermittent load shedding, having a backup generator that is only for the radiotherapy department has often proved to be of good use.” and ”[…] our main hazard is heavy rain and pouring water as well as pausing public traffic in impeding arrival of staff and patients.”, give an insight into the reality of global RT continuity problems and confirm the need for our research work. Finally, one participant mentioned many regional differences in treatment techniques due to local protocols. Concluding, it was also proposed that medical device manufacturers work towards international standardisation of treatment techniques, thereby supporting global risk mitigation.

#### Conceptual model

In consideration of the risk-minimising measures with high approval ratings, we present a conceptual model for natural disaster resilient RT for visualisation purposes and to further abstract our results, thus making them more accessible and applicable. The model shown in Fig 6 presents a treatment centre, a partner centre, patients and the relations between the different entities. Certain measures are applied within the centre, while others involve partners and patients. The model is based on a technology-intensive patient treatment that takes place in a time-defined sequence of appointments and in which practitioners have limited flexibility in the treatment programme. Therefore, the model could possibly also reflect other forms of therapy in healthcare.

**Fig 6.**
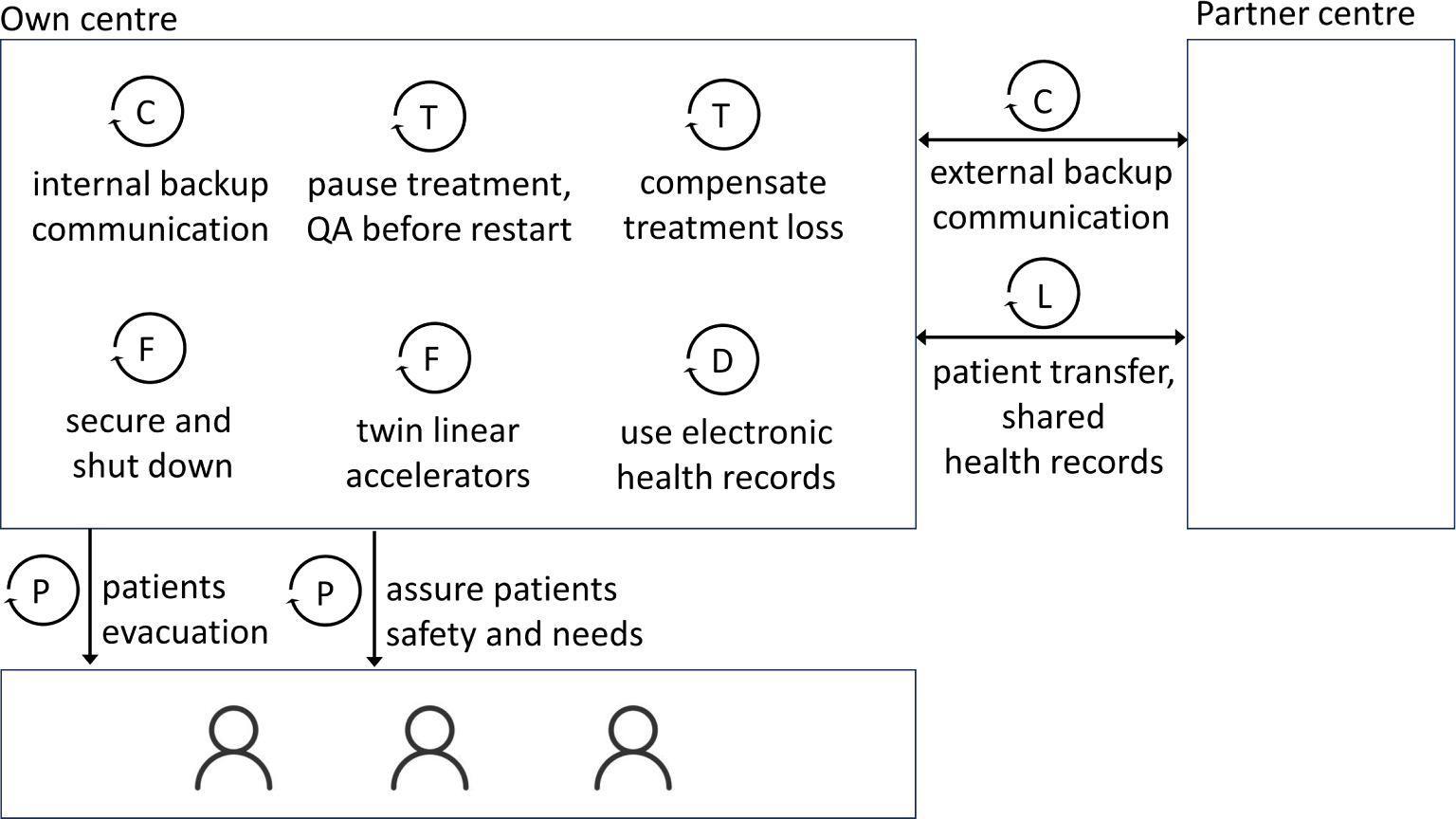
Conceptual model for natural disaster resilient RT. C: Communication theme, T: Treatment, F: Facility, D: Data theme, P: Patients, L: Collaboration:

## Discussion

### Validation

The Global Risk Report 2024 of the World Economic Forum presents findings from the Global Risks Perception Survey, incorporating insights from 1,500 experts worldwide. According to the survey, environmental risks are prominent in short and long-term outlooks. Notably, two-thirds of the experts identified extreme weather as the primary risk of causing a global crisis, underscoring the need for risk management strategies in addressing environmental challenges [42]. The figures confirm the risk assessment of the RT experts for the time period addressed in our research.

Additionally, the results of our RT research in the context of natural disasters are consistent with the findings for oncology care in general. For example, Saghir et al. identified areas of crucial consideration. In disaster-prone areas, ensuring healthcare resilience involves disseminating facility information through radio broadcasts, cloud-based social media, or satellite internet, which validates the *Communication* theme. According to Saghir et al., storing medical records in the cloud ensures treatment continuity, a finding that supports the *Data* theme of our taxonomy [43].

The WHO recommends adhering to building codes in high-risk areas to safeguard healthcare facilities. They also suggest cooperation between centres and medical associations to facilitate patient transfer and disaster response. However, rapid-response teams require specialised RT knowledge and familiarity with on-site equipment and processes [44]. The importance of such support is underlined by the strategic goal of the European Federation of Organisations for Medical Physics to develop a website-based portal to gather medical physics volunteers willing to assist in tackling health challenges [45]. Thus, the taxonomy’s themes *Facility* and *Collaboration* are validated.

Our findings also confirm a literature review by Ginex et al., who explored the impact of climate disasters on oncology care, including patients, healthcare staff, and the health systems. Patients faced treatment and communication disruptions, while the workforce was not prepared for disasters and experienced distress. Health systems encountered closures and shifted services, emphasising the need for improved emergency response plans. The study confirms that a holistic approach is necessary, including interventions to mitigate care interruptions, enhance coordination and planning, and improve resource allocation [46]. It validates primarily the themes *Organisation* and *Collaboration*.

Guzman and Malik highlight the importance of disaster management education programs for patients and clinicians. They ground their work on a literature review regarding cancer patients during and after disasters and conclude that patients face challenges, including limited assistance for chronic diseases, lack of medical history, and communication barriers [7], thereby validating mainly the *Organisation* and *Communication* themes of the taxonomy.

In addition, individual risk mitigation measures are confirmed by various academic publications. Communication among and towards professionals is an important aspect of risk mitigation, as reflected in the *Communication* theme. However, focus group analysis with RTTs in Sweden has revealed that communication among professional groups needs improvement even in non-disaster times. Therefore, the professional group of RTTs should be given appropriate attention in disaster preparation [47]. O’Sullivan-Steben et al. presented an RT patient portal accessible on mobile phones, providing patients with easy-to-understand cancer treatment data. They used the Minimal Common Oncology Data Elements (mCODE) data standard for treatment summaries that are easily shareable [48]. The approach describes a specific risk-mitigation solution and validates the *Data* theme. Dosanjh et al. elaborate on developing medical LINACs for challenging environments to provide robust, affordable, low-cost devices with corresponding IT solutions for Low and Middle-Income Countries. Many of the solutions presented, such as reducing dependence on clean water and stable energy as well as extended remote service options, increase the resilience of LINACs and would also be advantageous in times of crisis. In doing so, they primarily support the *Facility* and *Therapy* themes of our taxonomy [49]. Specific therapy options such as hypofractionation, the delivery of higher doses in fewer treatment fractions, or flexible therapy courses are suggested by Yom and Harari for head and neck cancer patients and thus confirm the *Therapy* theme [50]. A statement from Kovalchuk et al. also shows the importance of testing risk-mitigation measures before use: ”To continue treating our patients, we tried to connect the generator, which proved to be very challenging, as the simultaneous gantry rotation with the compressor was very power consuming” [51]. The statement from the field confirms the *Facility* and *Organisation* themes of our taxonomy.

Finally, a survey of California radiation oncologists evaluated emergency preparedness and the effect of wildfires on RT delivery, mapping the geographic distribution of wildfires to RT centre locations. The survey confirms the negative impact of natural disasters on RT practice, including evacuations, transportation interruptions, cancelling and rescheduling appointments, patient transfer, and physical, mental, or financial stress. Less than half of the RT centres had an emergency preparedness plan in place [**?**]. The survey confirms the themes *Protection*, *Access* and *Communication*.

In conclusion, the studies validate the different themes of the taxonomy presented in Fig 2. They also support the theoretical framework and underscore the significance of our research work.

### Reflection

This paper complements a series of publications on BCM in RT practice that support the effective delivery of therapy. In our previous research, we have analysed the influence of the COVID-19 pandemic on RT. We developed an intuitive decision support tool that informs about risk mitigation measures during pandemic times. The system uses a taxonomy based on global risk mitigation best-practices [9]. During the investigations of the influence of natural disasters on RT, it became apparent that the taxonomy could be extended to cope with the risks of natural disasters. The taxonomy presented in Fig 2 thus represents a meaningful extension.

By 2040, around 50 % of new cancer cases worldwide will occur in individuals aged 70 years and older [18]. Given this demographic trend, it becomes crucial to address the unique circumstances elderly individuals face during natural disasters. Older adults are at a higher risk of mortality, injuries, inadequate support from authorities, and post-disaster health issues. They also often face limitations or disabilities in vision, hearing, mobility, communication, and cognitive abilities [53]. Particularly for early-stage cancers, the lack of access to cancer care is notably linked to insufficient transportation options caused by natural disasters [54]. New treatment methods, such as hypofractionation, reduce the duration of the overall therapy and, thereby, its vulnerability to natural disasters. However, these methods require specialised equipment and skills, increasing business continuity vulnerability [55].

Furthermore, natural disasters are distributed differently worldwide and require regional or local strategies [41, 56]. In particular, rural areas face different challenges compared to urban areas and therefore require special attention [57]. Overall, BCM strategies must consider the different impacts of natural disasters on countries depending on their population density and development status [58]. Finally, man-made risks must also be considered when developing a concept for comprehensive RT BCM.

### Man-made threats

In addition to the influences of pandemics and natural disasters, man-made hazards should be considered in BCM. Cyberattacks severely impact healthcare delivery, sometimes with similar effects to those caused by natural disasters. After a cyberattack incident, the Radiation Oncology Division at the University of Vermont, for example, reported that patient information and schedules had to be reconstructed from paper records, and a triage system facilitated immediate treatment transfers. Medical physics and the IT department collaborated to restore services without backups or network connectivity. Treatments resumed incrementally as systems were rebuilt. The authors recommend individualised contingency plans based on centre-specific vulnerabilities, risks, and resources [59]. A 12-day interruption at the National University of Ireland Galway due to a cyberattack emphasised the importance of enhancing treatment compensation plans [60]. Also, the University of Maryland’s Department of Radiation Oncology reported an incident and consequently developed a solution to ensure treatment continuity during a cyberattack on the radiation oncology information system (ROIS) by automatically saving data to a secure server [61].

Finally, war and terrorism can lead to comparable results as natural disasters. The finely tuned RT processes are disturbed or interrupted, which is shown by a report from the war against Ukraine [51]. A negative side effect is the pressure on health systems in neighbouring states caused by war refugees [62].

### Limitations

There are limitations to our work. Despite the extensive scope of the SLR in terms of time and content, it cannot be excluded that relevant academic literature was published through other platforms. Due to the practical nature of BCM and RM, it can also not be ruled out that best practices were published in non-academic media. It could even be that RT centres did not want to publicise their experience or did not have the experience or ressources to do so. The SLR was performed by two independent researchers using a literature management tool to reduce selection bias and increase the quality of the results. Still, it is possible that relevant literature was sorted out if the title or abstract was not formulated specifically enough. While valuable for qualitative insights, manual content analysis presents limitations, including confirmation bias. Combining manual analysis with automated techniques could enhance the robustness and efficiency of future research. An independent additional review of our analysis results would further increase rigour.

Also, our online survey has certain limitations. Sampling bias could have occurred as the survey was publically accessible. The social media posts with the call to action to participate in the survey were not advertised in paid campaigns. The organic distribution based on social media algorithms likely resulted in higher impressions on first and second-level contacts. In future surveys, paid campaigns could be used to better address the target groups based on socio-demographic parameters, increase participation levels and be able to back up the analysis with campaign metrics. It must also be noted that the platforms and tools are not accessible in all countries due to Internet restrictions.

Finally, the research results are based on the opinions of RT specialists, while patients were not included in the survey. In the future, a patient study could add an additional perspective.

## Conclusion

This paper presents an SLR, content analysis, and confirmative online survey focusing on RT at risk caused by natural disasters. We also created a link to additional disruptions, such as pandemics and man-made disasters. Cancer treatment facilities are called to ensure disaster preparedness to withstand climate threats and should evaluate and mitigate their contributions to greenhouse gas emissions [63]. This is particularly true for RT practice with its technique intensity, dependency on skilled personnel, collaboration with co-therapists, and many treatment appointments.

As we collected subjective opinions of the experts with our survey, we plan to extent our investigation in a next step, starting in Germany, to get additional insights into the actual disaster preparednes of (German) radiotherapy centres. This will help us to further concretise and evaluate the risk mitigation measures. Following the international survey, a cross-cultural comparison in (objective and subjective) disaster preparedness would be interesting. A study could investigate how cultural factors influence disaster preparedness and response and the perceived need to do so. This research could help adapt disaster response strategies to match cultural nuances.

In previous research, we have developed a chatbot-based decision support system (DSS) for RT that suggests risk mitigation measures in pandemics [9]. In future work, we plan to expand the knowledge base of the chatbot with the accumulated knowledge of natural disaster risk mitigation. We also plan to integrate a tool that allows users to self-assess their degree of risk preparation. Both will help users to make informed decisions. We aim to make the DSS useful on the strategic and operational levels by including an adaptive component based on both the knowledge base and mathematical optimisation and simulation that offers real-time decision support during a crisis and a training component to increase disaster preparedness.

Performing an economic impact analysis of disaster response strategies to keep an RT practice going during a disaster would be another avenue for future research. A cost-effective analysis of various disaster response and preparedness initiatives could be performed.

Most cancer studies in disaster planning focus on hurricanes, floods, and earthquakes. Future research could examine the specific characteristics of other natural disasters on RT practice and evaluate the feasibility and cost of targeted solutions, such as mobile imaging and treatment solutions [64]. The difference between subjective and objective disaster preparedness should also be analysed using our conceptual model.

As stated in [70]: ”[…] it is predicted that by 2050 there will be 534 000 climate-related deaths worldwide […]”. The foreseen increase in cancer incidences in the coming decades reflects the importance of natural hazard research for RT treatment [71].

To address the vulnerability to natural hazards, it is crucial to consider all three components of vulnerability: exposure, sensitivity, and resilience. Strengthening collaboration between the climate change adaptation, environmental management, and poverty reduction communities and enhancing information exchange among these communities are necessary to reduce vulnerabilities effectively [72].

Part of this publication was written in the hot summer of 2023 in Greece while fueled by strong winds and temperatures up to 46 degrees Celsius (114.8*^◦^*F), many wildfires were out of control and in close local proximity [65, 66]. The wildfires being declared the largest ever recorded in the European Union [67] were followed by catastrophic rain resulting in landslides, road and bridge collapse, and severing water supplies [68]. Overall, this situation vividly illustrates the need for increased disaster preparedness efforts. As stated in the editorial of the International Journal of Radiation Oncology [69] ”The greatest threat facing us may be a failure of the imagination - failing to recognise the dangers around us as proximate realities, failing to combine preparedness, resourcefulness, and compassion to be ready for the disasters that could befall us, and others.”

## Data Availability

The survey results are included in the paper. The original survey data cannot be published online, but could be made available based on individual requests to the authors.

## Acknowledgments

The authors sincerely thank the survey participants who supported global development and dissemination of knowledge in times of high workload and responsibility. The authors would particularly like to thank the participants from war or crisis zones who, despite all difficulties, supported academic knowledge building.

**S1 Supporting information 1. Literature summary.**

**S2 Supporting information 2. Coding data.**

**S3 Supporting information 3. Survey data.**

**S4 Supporting information 4. Structured online questionnaire.**

## References

1. World Meteorological Organization. Provisional State of the Global Climate 2023. [accessed 2024 Mar 6] Available from URL: https://wmo.int/files/provisional-state-of-global-climate-2023.

2. Centre for Research on the Epidemiology of Disasters - CRED. 2023 Disasters in Numbers: A Significant Year of Disaster Impact. [accessed 2024 May 12] Available from URL: https://files.emdat.be/reports/2023EMDAT report.pdf

3. Centre for Research on the Epidemiology of Disasters. EM-DAT The International Disaster Database. [accessed 2024 Apr 21] Available from URL: https://www.emdat.be/.

4. United Nations Office for Disaster Risk Reduction. Sendai framework for disaster risk reduction 2015 - 2030. [accessed 2024 Mar 6] Available from URL: https://www.undrr.org/publication/sendai-framework-disaster-risk-reduction-2015-2030.

5. World Meteorological Organization. Bulletin. Early Warnings for All; 2023 72 (1). [accessed 2024 Mar 6] Available from URL: https://library.wmo.int/viewer/64131/download?file=WMO_Bulletin_72_1_en.pdf&type=pdf&navigator=1.

6. Romanello M, Di Napoli C, Drummond P, Green C, Kennard H, Lampard P et al. The 2022 report of the Lancet Countdown on health and climate change: health at the mercy of fossil fuels. Lancet 2022; 400(10363):1619–54.

7. Guzman R de, Malik M. Global Cancer Burden and Natural Disasters: A Focus on Asia’s Vulnerability, Resilience Building, and Impact on Cancer Care. Journal of Global Oncology 2019; 5:1–8.

8. Gorji HA, Jafari H, Heidari M, Seifi B. Cancer patients during and after natural and man-made disasters: a systematic review. Asian Pac J Cancer Prev 2018; 19:2695.

9. Reuter-Oppermann M, Müller-Polyzou R, Georgiadis A. Towards a decision support system for radiotherapy business continuity in a pandemic crisis. Journal of Decision Systems 2022; 31(1-2):40–67.

10. World Meteorological Organization. Natural hazards and disaster risk reduction; 2015 [accessed 2024 Mar 6]. Available from: URL: https://public.wmo.int/en/our-mandate/focus-areas/natural-hazards-and-disaster-risk-reduction.

11. United Nations Office for Disaster Risk Reduction. Sendai Framework Terminology on Disaster Risk Reduction: Hazard. 2023 [accessed 2024 Mar 6]. Available from: URL: https://www.undrr.org/terminology/hazard.

12. Bokwa A. Encyclopedia of natural hazards: Includes case studies 2013. [accessed 2024 Mar 6] Available from: URL: http://www.loc.gov/catdir/enhancements/fy1506/2012944445-d.html.

13. Chowdhury MAB, Fiore AJ, Cohen SA, Wheatley C, Wheatley B, Balakrishnan MP et al. Health Impact of Hurricanes Irma and Maria on St Thomas and St John, US Virgin Islands, 2017-2018. American Journal of Public Health 2019; 109(12):1725–32.

14. Kidder T. Recovering from disaster–Partners in Health and the Haitian earthquake. N Engl J Med 2010; 362(9):769–72.

15. Ryan BJ, Franklin RC, Burkle FM, Watt K, Aitken P, Smith EC, et al. Defining, Describing, and Categorizing Public Health Infrastructure Priorities for Tropical Cyclone, Flood, Storm, Tornado, and Tsunami-Related Disasters. Disaster Medicine and Public Health Preparedness 2016; 10(4):598–610.

16. Content AM, Staff N. How to Get Ready for a Natural Disaster When You Have Cancer [accessed 2023 Jun 30]. Available from: URL: https://www.cancer.org/cancer/latest-news/how-to-prepare-for-a-weather-emergency-when-you-have-cancer.html.

17. World Health Organization. Cancer; 2023 [accessed 2023 Jul 8]. Available from: URL: https://www.who.int/news-room/fact-sheets/detail/cancer.

18. The International Agency for Research on Cancer (IARC). Global Cancer Observatory; 2023 [accessed 2023 Jul 8]. Available from: URL: https://gco.iarc.fr/.

19. Atun R, Jaffray DA, Barton MB, Bray F, Baumann M, Vikram B et al. Expanding global access to radiotherapy. The Lancet Oncology 2015; 16(10):1153–86.

20. Müller-Polyzou R, Reuter-Oppermann M, Engbert A, Schmidt R. Identifying user assistance systems for radiotherapy to increase efficiency and help saving lives. Health Systems 2020:1–19.

21. ISO. ISO 22301:2019: Security and resilience, Business continuity management systems, Requirements; 2023 [accessed 2023 Jul 8]. Available from: URL: https://www.iso.org/standard/75106.html.

22. Webster J, Watson RT. Analyzing the Past to Prepare for the Future: Writing a Literature Review. MIS Quarterly 2002; 26(2):xiii–xxiii. Available from: URL: http://www.jstor.org/stable/4132319.

23. Gough D, Thomas J, Oliver S. Clarifying differences between review designs and methods. Syst Rev 2012; 1(1):28. Available from: URL: https://systematicreviewsjournal.biomedcentral.com/articles/10.1186/2046-4053-1-28.

24. Schlegel W, Karger CP, Jäkel O. Medizinische Physik: Grundlagen - Bildgebung - Therapie - Technik. 1. Aufl. 2018. Berlin, Heidelberg: Springer Spektrum; 2018.

25. Citavi: Best Reference Management Software for Writing and Note Taking. 2023 [accessed 2023 Jun 25]. Available from: URL: https://www.citavi.com/en.

26. Liberati A, Altman DG, Tetzlaff J, Mulrow C, Gøtzsche PC, Ioannidis JPA et al. The PRISMA statement for reporting systematic reviews and meta-analyses of studies that evaluate healthcare interventions: explanation and elaboration. BMJ 2009; 339:b2700.

27. Google Forms: Online form creator [Google Workspace]; 2023 [accessed 2023 Nov 4]. Available from: URL: https://www.google.com/intl/en-GB/forms/about/.

28. Sullivan GM, Artino AR. Analyzing and interpreting data from likert-type scales. Journal of Graduate Medical Education 5(4):541–2.

29. Anacak Y, Kurtul N, Nasuhbeyŏglu D, Oymak E, Ö nal HC. Radiotherapy facilities after the Türkiye-Syria earthquakes: lessons from the tragedy. The Lancet Oncology 2023; 24(4):312–4. Available from: URL: https://www.thelancet.com/journals/lanonc/article/piis1470-2045(23)00099-2/fulltext.

30. Espinel Z, Nogueira LM, Gay HA, Bryant JM, Hamilton W, Trapido EJ et al. Climate-driven Atlantic hurricanes create complex challenges for cancer care. The Lancet Oncology 2022; 23(12):1497–8. Available from: URL: https://www.thelancet.com/journals/lanonc/article/piis1470-2045(22)00635-0/fulltext.

31. Gay HA, Santiago R, Gil B, Remedios C, Montes PJ, Ĺopez-Araujo J, et al. Lessons Learned From Hurricane Maria in Puerto Rico: Practical Measures to Mitigate the Impact of a Catastrophic Natural Disaster on Radiation Oncology Patients. Practical Radiation Oncology 2019; 9(5):305–21. Available from: URL: https://www.sciencedirect.com/science/article/pii/s1879850019300797.

32. Grew D, Vatner R, DeWyngaert K, Nicholas S, Formenti S. The Impact of Superstorm Sandy on the Care of Radiation Oncology Patients. International Journal of Radiation Oncology Biology Physics 2013; 87(2):S490–S491.

33. Joob B, Wiwanitkit V. Lesson for management of cancerous patient in the big flooding. Journal of Cancer Research and Therapeutics 2012; 8(1):165–6. Available from: URL: https://journals.lww.com/cancerjournal/fulltext/2012/08010/lessonformanagementofcancerouspatientinthe.45.aspx.

34. Lopez-Araujo J, Burnett OL. Letter from Puerto Rico: The State of Radiation Oncology After Maria’s Landfall. International Journal of Radiation Oncology, Biology, Physics 2017; 99(5):1071–2.

35. Man RX-G, Lack DA, Wyatt CE, Murray V. The effect of natural disasters on cancer care: a systematic review. The Lancet Oncology 2018; 19(9):e482–e499.

36. Mireles M, Pino R, Teh BS, Farach A, Joseph A, Butler EB. Radiation Oncology in the Face of Natural Disaster: The Experience of Houston Methodist Hospital. International Journal of Radiation Oncology, Biology, Physics 2018; 100(4):843–4.

37. Ozaki A, Tsubokura M. Radiation Oncology and Related Oncology Fields in the Face of the 2011 ”Triple Disaster” in Fukushima, Japan. International Journal of Radiation Oncology, Biology, Physics 2018; 100(4):845–8.

38. Pérez-Andújar A. Puerto Rico: After Maŕıa. International Journal of Radiation Oncology, Biology, Physics 2018; 100(4):834–5.

39. Roach MC, Robinson CG, Bradley JD. Natural Disasters and the Importance of Minimizing Subsequent Radiation Therapy Interruptions for Locally Advanced Lung Cancer. International Journal of Radiation Oncology, Biology, Physics 2018; 100(4):836–7.

40. Royce TJ, Papagikos MA, Maguire PD, Marks LB. Carolina Hurricanes. International Journal of Radiation Oncology, Biology, Physics 2019; 103(3):775–6.

41. Ritchie H, Rosado P, Roser M. Natural Disasters; 2022. Available from: URL: https://ourworldindata.org/natural-disasters.

42. World Economic Forum. The Global Risks Report 2024: 19th Edition [Insight Report] 2024.

43. El Saghir NS, Soto Pérez de Celis, Enrique, Fares JE, Sullivan R. Cancer Care for Refugees and Displaced Populations: Middle East Conflicts and Global Natural Disasters. American Society of Clinical Oncology Educational Book 2018; 38:433–40.

44. World Health Organization. Emergency medical teams; 2023 [accessed 2024 May 12]. Available from: URL: https://www.who.int/emergencies/partners/emergency-medical-teams.

45. Koutsouveli E. Incoming President of the European Federation of Organizations for Medical Physics editorial. Phys Med 2024; 117:103197.

46. Ginex P, Dickman E, Elia MR, Burbage D, Wilson R, Koos JA et al. Climate disasters and oncology care: a systematic review of effects on patients, healthcare professionals, and health systems. Support Care Cancer 2023; 31(7):403.

47. Widmark C, Tishelman C, Gustafsson H, Sharp L. Information on the fly: Challenges in professional communication in high technological nursing. A focus group study from a radiotherapy department in Sweden. BMC Nursing 2012; 11:10.

48. O’Sullivan-Steben K, Galarneau L, Judd S, Laizner AM, Williams T, Kildea J. Design and implementation of a prototype radiotherapy menu in a patient portal. Journal of Applied Clinical Medical Physics 2024; 25(3):e14201.

49. Dosanjh M, Aggarwal A, Pistenmaa D, Amankwaa-Frempong E, Angal-Kalinin D, Boogert S, et al. Developing Innovative, Robust and Affordable Medical Linear Accelerators for Challenging Environments. Clin Oncol (R Coll Radiol) 2019; 31(6):352–5.

50. Yom SS, Harari PM. When Disaster Strikes: Mitigating the Adverse Impact on Head and Neck Cancer Patients. International Journal of Radiation Oncology, Biology, Physics 2018; 100(4):838–40.

51. Kovalchuk N, Zelinskyi R, Hanych A, Severyn Y, Bachynska B, Beznosenko A et al. Radiation Therapy Under the Falling Bombs: A Tale of 2 Ukrainian Cancer centres. Advances in Radiation Oncology 2022; 7(6):101027. Available from: URL: https://www.advancesradonc.org/article/S2452-1094(22)00133-6/fulltext.

52. Lichter KE, Baniel CC, Do I, Medhat Y, Avula V, Nogueira LM et al. Effects of Wildfire Events on California Radiation Oncology Clinics and Patients. Advances in Radiation Oncology 2024; 9(3):101395.

53. Maltais D. Elderly People With Disabilities And Natural Disasters: Vulnerability Of Seniors And Post Trauma. HSOA Journal of gerontology & geriatric medicine 2019; 5(4):1–7. Available from: URL: https://constellation.uqac.ca/id/eprint/5840/.

54. Loehn B, Pou AM, Nuss DW, Tenney J, McWhorter A, DiLeo M et al. Factors affecting access to head and neck cancer care after a natural disaster: a post-Hurricane Katrina survey. Head and Neck 2011; 33(1):37–44.

55. van Dyk J, editor. The modern technology of radiation oncology: A compendium for medical physicists and radiation oncologists. Madison, Wis.: Medical Physics Pub; 2020.

56. Hannah Ritchie and Pablo Rosado. Our World in Data. Natural Disasters [accessed 2024 Mar 3]. Available from: URL: https://ourworldindata.org/natural-disasters.

57. Chan EYY, Man AYT, Lam HCY. Scientific evidence on natural disasters and health emergency and disaster risk management in Asian rural-based area. British Medical Bulletin 2019; 129(1):91–105.

58. Donatti CI, Nicholas K, Fedele G, Delforge D, Speybroeck N, Moraga P et al. Global hotspots of climate-related disasters. International Journal of Disaster Risk Reduction 2024; 108:104488. Available from: URL: https://www.sciencedirect.com/science/article/pii/S2212420924002504.

59. Nelson CJ, Soisson ET, Li PC, Lester-Coll NH, Gagne H, Deeley MA et al. Impact of and Response to Cyberattacks in Radiation Oncology. Advances in Radiation Oncology 2022; 7(5):100897.

60. O’Shea K, Coleman L, Fahy L, Kleefeld C, Foley MJ, Moore M. Compensation for radiotherapy treatment interruptions due to a cyberattack: An isoeffective DVH-based dose compensation decision tool. Journal of Applied Clinical Medical Physics 2022; 23(9):e13716.

61. Zhang B, Chen S, Nichols E, D’Souza W, Prado K, Yi B. A practical cyberattack contingency plan for radiation oncology. Journal of Applied Clinical Medical Physics 2020; 21(7):181–6.

62. Wareńczak-Florczak Ż, Urbański B. What challenges does the humanitarian crisis and large number of refugees from Ukraine pose for Polish oncology? Reports of Practical Oncology and Radiotherapy 2022; 27(3):566–70. Available from: URL: https://journals.viamedica.pl/rpor/article/view/90020.

63. Nogueira LM, Yabroff KR, Bernstein A. Climate change and cancer. CA Cancer Journal for Clinicians 2020; 70(4):239–44.

64. Lung Foundation Australia. World First Mobile Screening Truck at Parliament Proves How Easy Saving Lives Could Be - Lung Foundation Australia; 2023 [accessed 2023 Nov 5]. Available from: URL: https://lungfoundation.com.au/news/world-first-mobile-screening-truck-at-parliament-proves-how-easy-saving-lives-could-be/.

65. Keep Talking Greece Fire in Velestino out of control, heading to Volos; one firefighter injured; 2023 [accessed 2023 Jul 30]. Available from: URL: https://www.keeptalkinggreece.com/2023/07/26/fire-velestino-volos-out-of-control/.

66. The Guardian Wildfire damages ammunition depot near Greek city of Volos – video; 2023 [accessed 2023 Jul 30]. Available from: URL: https://www.theguardian.com/world/video/2023/jul/28/wildfire-damages-ammunition-depot-near-greek-city-of-volos-video.

67. The Guardian Greece wildfire declared largest ever recorded in EU. The Guardian 2023 Aug 29 [accessed 2023 Sep 10]. Available from: URL: https://www.theguardian.com/world/2023/aug/29/greece-wildfire-declared-largest-ever-recorded-in-eu.

68. The Guardian. A biblical catastrophe: death toll rises to four as Storm Daniel lashes Greece. The Guardian 2023 Sep 7 [accessed 2023 Sep 10]. Available from: URL: https://www.theguardian.com/world/2023/sep/07/a-biblical-catastrophe-death-toll-rises-to-four-as-storm-daniel-lashes-greece.

69. Yom SS, Zietman AL. Radiation Therapy in a Time of Disaster. International Journal of Radiation Oncology, Biology, Physics 2018; 100(4):832–3. Available from: URL: https://www.osti.gov/biblio/23082803.

70. The Lancet Oncology. Editorial. Climate crisis and cancer: the need for urgent action. Lancet Oncol 2021; 22(10):1341.

71. Bray F, Laversanne M, Sung H, Ferlay J, Siegel RL, Soerjomataram I, et al. Global cancer statistics 2022: GLOBOCAN estimates of incidence and mortality worldwide for 36 cancers in 185 countries. CA: A Cancer Journal for Clinicians 2024.

72. Thomalla F, Downing T, Spanger-Siegfried E, Han G, Rockström J. Reducing hazard vulnerability: towards a common approach between disaster risk reduction and climate adaptation. Disasters 2006; 30(1):39–48.

